# Neonatal Seizure Detection Using Combined aEEG and Compressed Spectral Array Features: A Machine-Learning Proof-of-Concept Study

**DOI:** 10.64898/2026.07.02.26354953

**Authors:** Sylvia Edoigiawerie, Julia Henry, Brett Beaulieu-Jones, Henry David, Naoum P. Issa

**Affiliations:** Biological Sciences Division, The University of Chicago Pritzker School of Medicine, Chicago, Illinois, USA; AdventHealth for Children, Pediatric Neurology, Orlando, Florida, USA; Department of Medicine, The University of Chicago Medicine, Chicago, Illinois, USA; Department of Pediatrics, The University of Chicago Medicine, Chicago, Illinois, USA; Department of Neurology, The University of Chicago Medicine, Chicago, Illinois, USA

**Author notes:** Corresponding author: Naoum P. Issa, Comprehensive Epilepsy Center, Department of Neurology, 5841 S. Maryland Ave., MC 2030, University of Chicago, Chicago, IL 60637.

**Keywords:** hypoxic ischemic encephalopathy, neonates, seizure detection, biomarkers, algorithm

## Abstract

**Background:** To build a clinically translatable neonatal seizure detection algorithm using amplitude-integrated electroencephalography (aEEG) and compressed spectral array (CSA).

**Methods:** Using a public dataset of annotated neonatal EEGs, features of the aEEG and CSA were extracted from the left and right centroparietal electrodes. These features were then used to train and test three machine learning classifiers, Random Forest (RF), Support Vector Machines (SVM), and Artificial Neural Networks (ANN).

**Results:** The trained RF, SVM, and ANN classifiers had areas under the curve (AUC) of 0.80, 0.69, and 0.79 for capturing seizure time periods and an average accuracy of 0.91, 0.90, and 0.92 respectively for capturing seizure and non-seizure time periods. Median accuracy scores were higher among patients without hypoxic-ischemic encephalopathy (HIE; median = 1 for all three classifiers) than HIE patients (median = 0.92, 0.93, 0.93).

**Conclusion:** A clinically interpretable aEEG-CSA algorithm is feasible for neonatal seizure detection by extracting standard EEG features and coupling these features with a supervised ML classifier.

## Introduction

Neonates have the highest risk of developing seizures among all age groups (1). According to the American Clinical Neurophysiologic Society, an electrographic seizure is characterized by a rhythmic, evolving, and stereotyped EEG pattern lasting at least 10 seconds, regardless of whether clinical signs are present (2). Since most neonatal seizures lack overt clinical manifestations, manual, continuous EEG monitoring is the gold standard for seizure detection (1,3,4). However, 24-hour EEG monitoring generates large volumes of data that require expert interpretation, and the limited availability of pediatric neurophysiologists contributes to delays in seizure identification (5).

To expedite EEG review, quantitative EEG (qEEG) tools compress and transform EEG signals to highlight patterns that may correspond to seizures (6,7). Among these, amplitude-integrated EEG (aEEG) and compressed spectral array (CSA) are widely used bedside trending tools in Neonatal Intensive Care Units. However, these tools are not substitutes for conventional EEG; instead, they are intended to support EEG interpretation, particularly when continuous expert review is not feasible (8).

aEEG is the most commonly used long-term trending tool for neonatal monitoring (9,10). The aEEG enables clinicians to detect time periods suspicious for seizure activity by identifying abrupt rises in the amplitude of the aEEG, also known as the upper and lower margins (11,12). However, manual aEEG interpretation has limited sensitivity: in a large multicenter study, neonatologists detected only 25% of electrographic seizures (13). In addition, aEEG performance varies across proprietary algorithms and still requires expert review (13,14). For these reasons, aEEG is best used as a supplementary trend rather than a standalone diagnostic tool (14).

CSA displays EEG power across frequency bands (0.5–30 Hz) as a compressed spectral plot. Clinicians can identify areas suspicious for seizure activity as a stereotyped “flame” spectrogram pattern, which reflects an increase in power across a range of frequencies that contrasts sharply with the background activity (15). While spectral analysis has demonstrated utility in adult seizure detection (16–19), its performance in neonates has been less extensively validated, and its integration with other qEEG features remains an area of active investigation. As with aEEG, CSA is most effective when integrated with conventional EEG interpretation, since it provides complementary frequency-based information that can help clinicians identify evolving electrographic patterns.

In this study, we extracted features from both aEEG and CSA to train supervised machine-learning (ML) classifiers for neonatal seizure detection. Combining these two qEEG modalities leverages their complementary strengths: aEEG provides amplitude-based trends, while CSA captures frequency-based changes. Both modalities show trends that often precede or accompany electrographic seizures. This combination mirrors how clinicians integrate multiple EEG trends at the bedside. ML classifiers were selected because they can model nonlinear relationships among multiple EEG features and have shown promise in doing so in prior neonatal seizure detection research (20–22). Our goal was to evaluate whether both aEEG and CSA features could support automated seizure detection in a transparent, clinically meaningful way. The aim of this work is to serve as a proof of concept that a combination of aEEG and CSA features can be used for detecting neonatal seizures and determining seizure burden.

## Methods

### Preprocessing

EEG data was obtained from a publicly available database of neonatal EEG recordings from Helsinki University (23). EEGs from 79 neonates, 39 of whom had at least one seizure, were sampled at 256 Hz. Of the various disease etiologies represented in the set of EEGs, Hypoxic Ischemic Encephalopathy (HIE) due to birth asphyxia constituted the largest subgroup with 35 patients, 24 of whom had seizures. Post-menstrual ages at the time of recording ranged from 34 to 45 weeks. Very preterm infants (less than 32 weeks) were excluded. Other available clinical data included were sex, birthweight, diagnosis, and a short summary of neuroimaging findings; however, no MRI files were made available (23). EEG recordings were annotated separately for seizures by three clinicians; annotations of seizure location were not included. Each recording was evaluated using the consensus (‘AND’) label across all three reviewers for training and testing as it provides the most stringent seizure classification. Using this consensus annotation, the dataset contains 302 total seizures. EEGs were bandpass filtered from 0.5 to 40 Hz using a 6-pole Butterworth filter and MATLAB function “filtfilt” for zero-phase distortion. An example EEG channel with consensus seizure periods marked from patient four is shown in Figure 1A. All data analysis was completed using MATLAB 2022b and GraphPad Prism.

**Figure 1.**
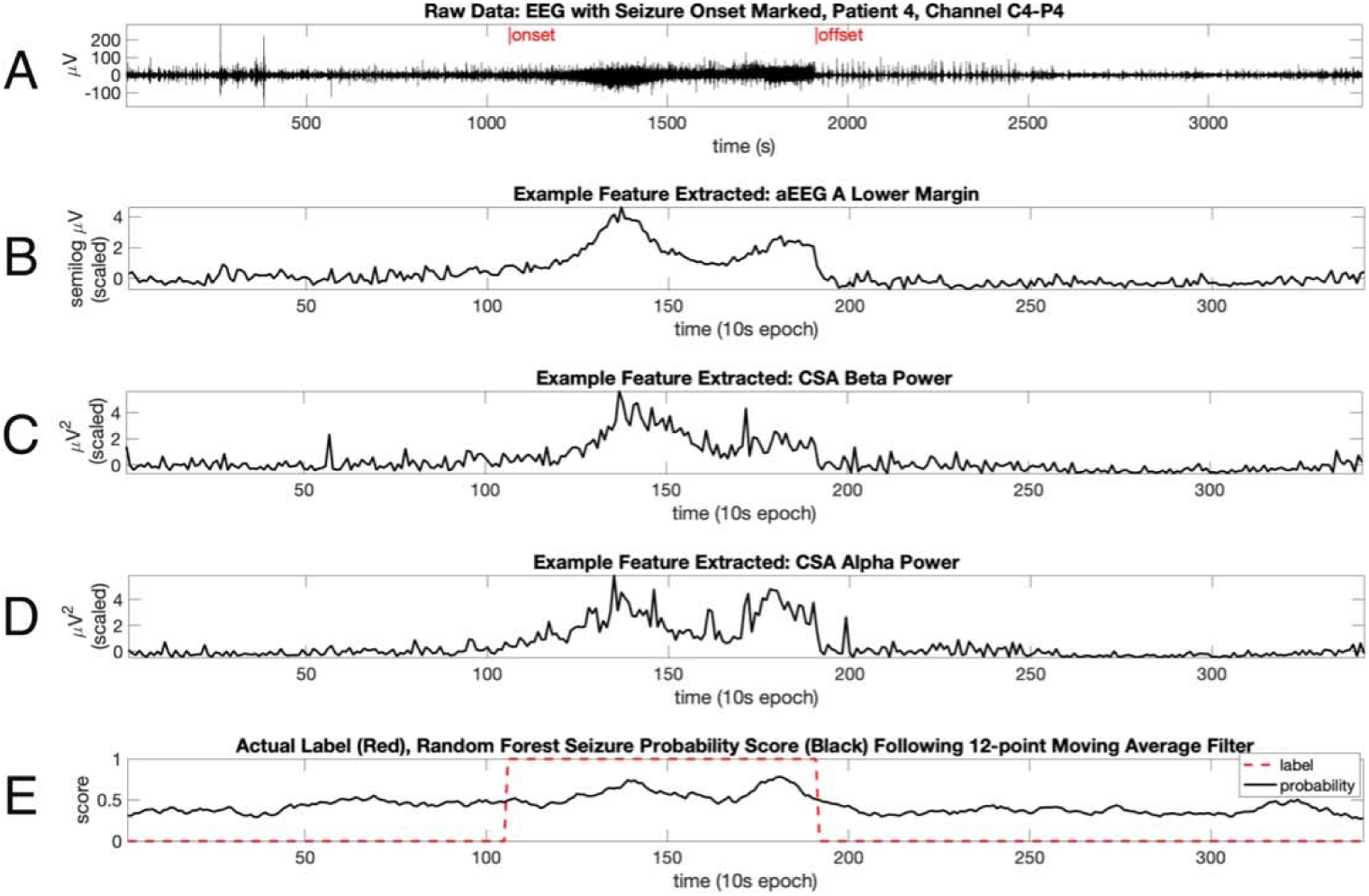
A. Raw EEG time series from channel C4–P4 with seizure onset and offset marked using the consensus annotation. B. The aEEG A lower margin demonstrates the characteristic upward shift in amplitude trend during seizure periods. C and D. CSA beta-band power (C) and alpha-band power (D) both show distinct increases during seizure windows, reflecting the frequency-based recruitment seen during electrographic seizure evolution. E. Random Forest classifier seizure probability scores for each 10-second epoch (black trace), shown alongside consensus seizure labels (red dashed line; 0 = non-seizure, 1 = seizure).

### Feature Extraction

Eleven features were extracted separately from the left and right centroparietal electrodes (C3-P3, C4-P4): six aEEG features and five CSA features. We focused on the bipolar C3-P3 and C4-P4 derivations because these centroparietal channels have demonstrated high sensitivity for neonatal seizure detection and are commonly used for generating bedside qEEG trends (13,24,25). We then segmented the EEG into non-overlapping 10-second windows for feature extraction. This window length aligns with the neonatal electrographic seizure definition, which requires an evolution pattern lasting at least 10 seconds (2). Using 10-second epochs ensures that each window can contain a complete electrographic seizure event while maintaining computational efficiency. Each 10-second epoch was represented by a vector of features from the aEEG and CSA.

#### aEEG Feature Extraction

The procedure of Ding and Zhang et al 2013 was adapted to compute the aEEG margins and aEEG envelope (26). Each EEG was filtered using an asymmetric bandpass filter (2-15 Hz), absolute-value rectified, and smoothed with a moving average filter (Fig. 2B). The EEG was then segmented into 10-second windows and the amplitude distribution in each window was calculated to extract the upper (93^rd^ percentile sample) and lower (9^th^ percentile sample) amplitude values. The amplitudes were converted into a semilogarithmic scale so amplitudes above 10 µV were log-transformed (base-10), while amplitudes below 10 µV were scaled by dividing by 10.

**Figure 2.**
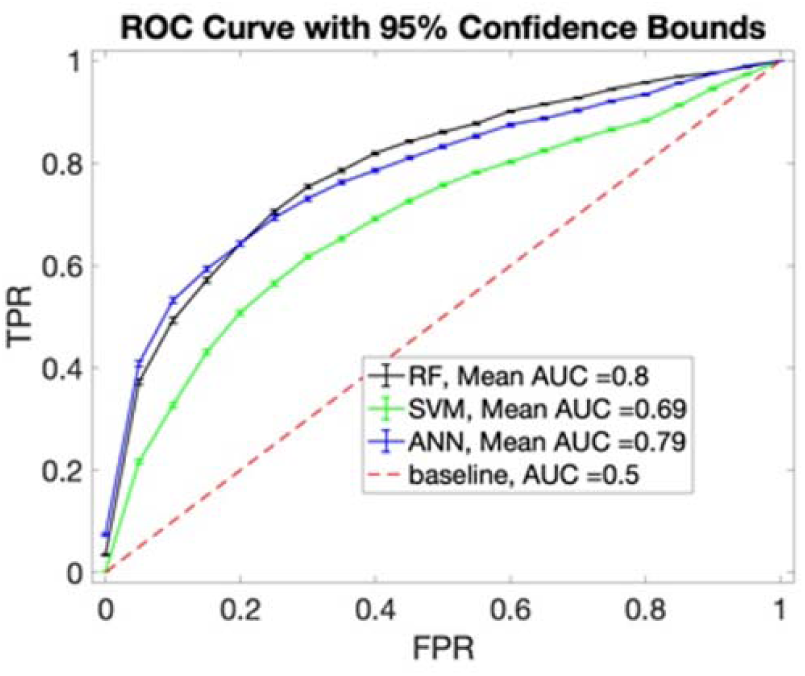
Performance of the aEEG–CSA seizure-detection algorithm across the Random Forest (RF), Support Vector Machine (SVM), and Artificial Neural Network (ANN) classifiers. The mean AUC values indicate that the RF classifier achieves the highest overall performance, followed by the ANN and SVM classifiers. The red dashed line marks chance-level performance (AUC = 0.5), which corresponds to random classification.

Two notable changes were made from the published protocol of Ding and Zhang 2013. One was increasing the temporal resolution of the aEEG. While Ding and Zhang 2013 computed the aEEG in 5-minute windows to mimic the resolution of the clinical aEEG (26), a 10-second window was chosen since it is better suited for capturing brief seizures and allows for finer temporal resolution. The second change was to generate two aEEGs, aEEG A and aEEG B, which differ in the duration of the moving average window used for smoothing (Supplemental Table 1). The two smoothing parameters were selected to reflect known variability in proprietary aEEG algorithms (27). Werther et al 2017 found that the smoothing filter parameters accounted for performance differences among proprietary aEEG systems (27). We directly adapted the filter parameters described by Werther et al 2017 for the Olympic and NicoletOne systems (27).

In total, six aEEG features were extracted for every 10-second EEG window: the upper margin, lower margin and median envelope using the aEEG A algorithm and aEEG B algorithm.

#### CSA Feature Extraction

To make a Compressed Spectral Array (CSA), the power spectra across each 10-second window of the preprocessed EEG was computed. From each spectrum, the power across each clinical frequency band, delta, theta, alpha, and beta, were computed from both channels. While delta activity is the dominant neonatal EEG rhythm (28), we included the full set of standard clinical frequency bands for completeness and to maintain consistency with conventional CSA displays used at the bedside. In addition, the spectral slope across the delta to beta (0.5-30Hz) was computed using Matlab’s “spectralSlope” function.

### Artifact Removal and Data Scaling

To reduce the effect of electrical and movement artifact on seizure detection, a heuristic artifact removal procedure was used. Any epoch with an aEEG lower margin value below 0.001µV or with total power below 0.001µV^2^ in the 0.5-30 Hz band was removed since these epochs corresponded to recording pauses in the Helsinki dataset (23). Also, any epoch with an upper margin or total power greater than five times the median value for the patient was removed to eliminate high-amplitude movement artifact. No epochs labeled as seizure were removed. Out of 40,251 total epochs, 916 artifact epochs were removed with 39,335 epochs remaining for training and testing. After artifact removal, each feature was scaled per patient using the median and interquartile range such that each feature had an interquartile range of one. This form of scaling was implemented since it is less affected by extreme outliers than methods that use mean and standard deviation (29).

### Synthetic Data, Model Training, and Testing

Patients were split into three groups for three-fold cross-validation using a stratified K-Fold approach to ensure the proportions of seizure and non-seizure patients was equivalent across all groups. Three-fold cross-validation was chosen because the dataset was relatively small. To address the substantial class imbalance, synthetic seizure samples were generated using the Adaptive Synthetic Oversampling Technique (ADASYN), which creates new minority-class samples in regions where seizure epochs are sparse by interpolating feature values from nearest-neighbor seizure epochs (30,31).

Training and testing were done using three supervised ML algorithms: Random Forest (RF), Support Vector Machine (SVM), and an Artificial Neural Network (ANN). These models were selected because they represent distinct learning paradigms, tree-based, margin-based, and neural-network–based approaches (32–34), which allows us to evaluate whether classifier architecture influences performance when using aEEG and CSA features. The RF classifier was generated using MATLAB’s “fitcensemble” function using the bagging method with all 11 feature variables selected for sampling per tree. An SVM classifier with a Radial Basis Function (RBF) kernel was applied using MATLAB’s “fitcsvm” function (35). SVM using an RBF kernel has been shown to be effective for neonatal seizure detection with a different feature set (20). Weighted standardization of predictors was used to improve performance (35). For the ANN, a simple feedforward neural network was implemented using MATLAB’s “fitcnet” classifier (36). The network consisted of two fully connected hidden layers, with a ReLu activation layer applied to the first fully connected layer and a SoftMax function applied to the final activation layer (36). Z-score feature standardization was selected via hyperparameter tuning. Finally, since neonatal seizures are often focal, separate left-sided (C3-P3) and right-sided (C4-P4) models were trained for each classifier.

### Post-processing and Algorithm Performance Assessments

For post-processing, seizure prediction probability scores were smoothed using a moving average filter. Performance was first evaluated using a range of smoothing window lengths to determine the effect of smoothing on the percentage of seizures detected and the false positive rate (FPR). AUC and accuracy scores were calculated using the 12-point window. This window length represents smoothing across 120 seconds, since each epoch represents 10-seconds of EEG. This window duration approximates the average seizure duration of 115 seconds in this dataset. The maximum filtered probability scores across both electrodes were then compared to consensus seizure labels (Figure 1E) and used to assess algorithm performance via receiver operating characteristic curve (ROC) analysis. Model performance was further evaluated using 1,000 bootstrap iterations, which generated confidence intervals for all performance metrics by repeatedly resampling the test-set predictions.

The optimal seizure probability threshold per classifier, AUC, and algorithm accuracy were computed using MATLAB’s “perfcurve” and MATLAB’s confusion matrix function “confusionmat.” Accuracy is calculated as the number of correctly identified epochs, whether true seizure or true non-seizure, divided by the total number of epochs. The AUC shows how well seizure and non-seizure time periods are classified at various seizure-probability thresholds for a given classifier.

Feature importance analysis was conducted using the RF Out-of-Bag permutation entropy via MATLAB’s “oobPermutedPredictorImportance” function (37,38). This method uses the out-of-bag error, which measures the RF classifier’s performance on samples not used to train each decision tree in the random forest. Features are scored by permuting each feature variable and assessing whether removing the feature increases or decreases the error. More important features cause greater increases in the error when removed. The mean importance score was taken between the two aEEGs.

## Results

### Algorithm Performance Across All Patients

The RF classifier performed the best of the three classifiers following bootstrapping with 1000 iterations (Figure 2A; mean AUC performance was 0.8, 0.69, and 0.79 for the RF, SVM and ANN classifiers respectively). The dataset contained 39,335 non-artifact epochs, and 4,224 of these epochs contained seizure activity.

### Feature Importance Performance for Seizure Classification

To determine which features contributed most to classification, we estimated importance scores for the RF classifier (Figure 3). The mean out-of-bag permutation importance scores were largest for CSA Beta power, the aEEG Lower Margin, and CSA alpha power (Figure 3). These three features, and their association with seizure activity, can also be seen in Figure 1 panels B, C, and D. As seizure activity begins there is a corresponding increase in each of these features.

**Figure 3.**
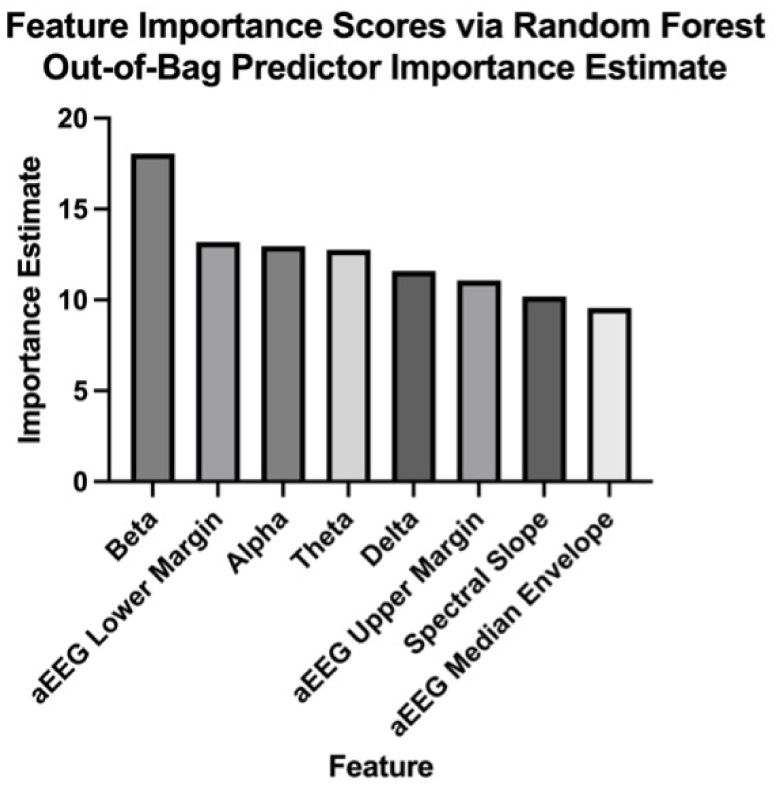
Importance scores for all 11 extracted aEEG and CSA features generated using the Random Forest out-of-bag predictor importance method. The rankings show that CSA beta power, the aEEG lower margin, and CSA alpha power were the most informative features for distinguishing seizure from non-seizure epochs.

### Per-Patient Performance Assessments

#### Per-Patient Accuracy Differs for Birth Asphyxia Patients

Algorithm performance was also assessed across individual patients. The median accuracy scores across all patients for the RF, SVM, and ANN classifiers were 0.99 (Figure 4A). To ascertain if seizure detection accuracy was different for patients with or without HIE, we applied multiple Wilcoxon signed rank tests that compared median accuracy between HIE and non-HIE patients for each classifier. P-values were corrected for multiple comparison using the Bonferroni-Dunn correction (Figure 4B). Infants with HIE due to birth asphyxia had significantly lower accuracy scores (median = 0.92, 0.93, 0.93 for RF, SVM, and ANN) than those without birth asphyxia (median = 1 for all three classifiers; p = 0.000018, 0.000063, and 0.000048 for the RF, SVM, and ANN).

**Figure 4.**
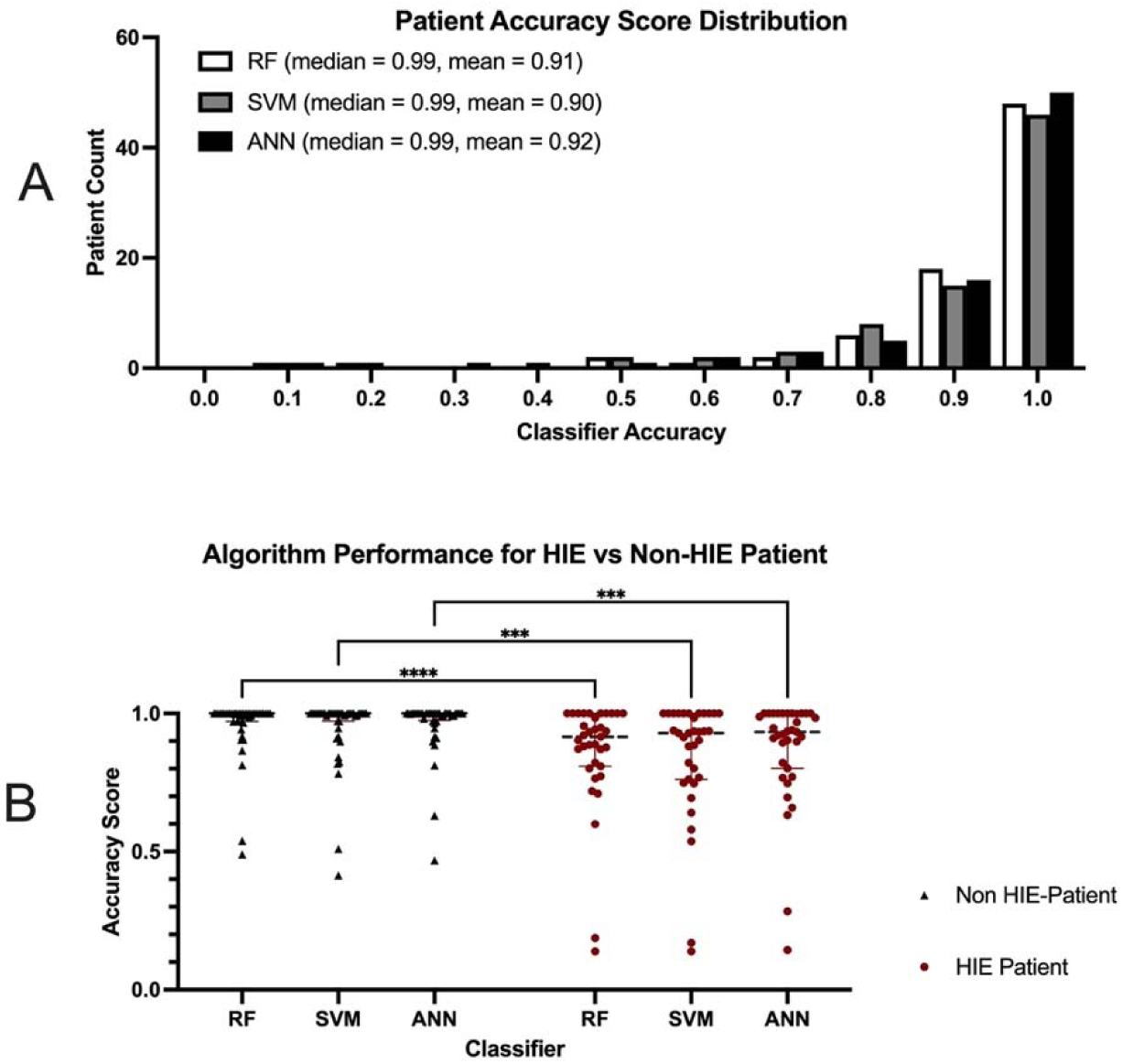
A. Accuracy scores for all 79 patients calculated using a single seizure-probability threshold applied uniformly across patients. Accuracy reflects the proportion of correctly classified epochs. B. Median accuracy values for each classifier grouped by disease etiology. Non-asphyxia patients are shown as black triangles (n = 44), and patients with HIE from birth asphyxia are shown as red circles (n = 35). HIE patients have lower median accuracy (0.92, 0.93, 0.93 for RF, SVM, and ANN) compared with non-HIE patients (median = 1 for all classifiers); P-values are 0.000018, 0.000063, and 0.000048 for RF, SVM, and ANN.

#### Effect of Smoothing on Algorithm Performance

We assessed the effect of smoothing the seizure probability scores during post-processing on the performance of the Random Forest Classifier. As expected, smoothing reduced the false positive rate (Figure 5A) but at the cost of reduced seizure detection rate (Figure 5B). The detection rate is highest (Figure 5C, window = 1, 82% of seizures detected) when no smoothing is applied, but this comes with the highest false positive rate (Figure 5C, FPR = 0.18).

**Figure 5.**
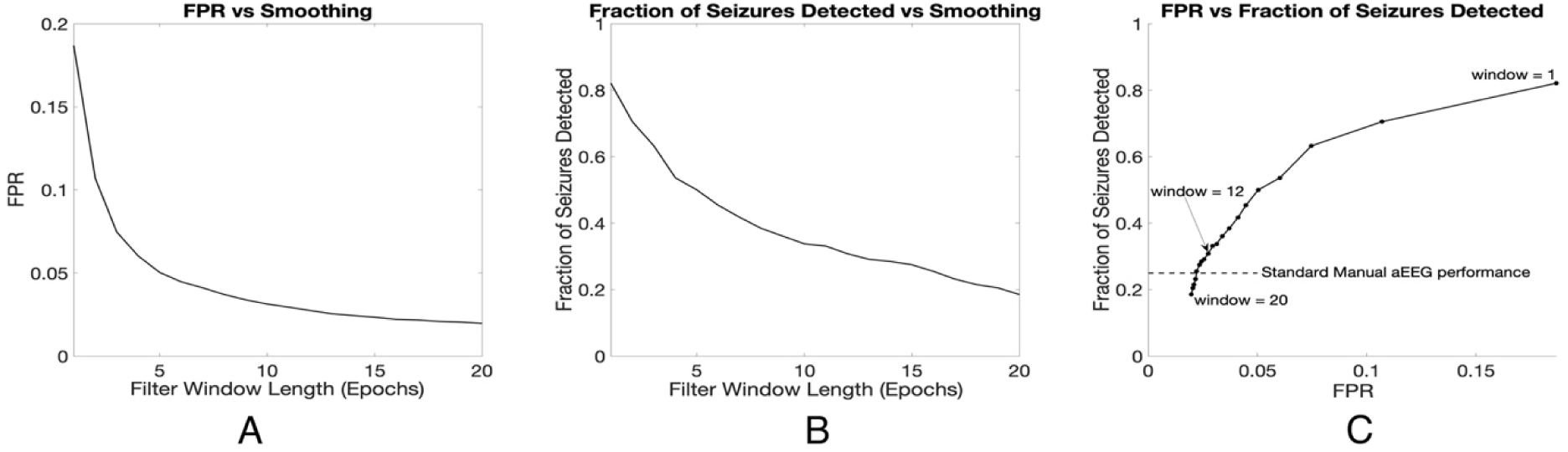
Tradeoff between sensitivity and false positive rate (FPR) as the seizure-probability scores are smoothed. A. Increasing the smoothing window lowers the FPR. B. Larger smoothing windows reduce the fraction of seizures detected. C. Smoothing affects both metrics simultaneously: as the window length increases, both FPR and seizure-detection rate decline. A 12-epoch window (120 seconds) was used in our analyses because it approximates the average seizure duration in this dataset (115 seconds). Smoothing windows longer than 16 epochs (160 seconds) produced detection rates below those reported for manual aEEG review (dashed line).

At the opposite extreme, heavy smoothing produces a low false positive rate (0-0.05), (Figure 5C), but also reduces sensitivity to 25%, which is comparable to manual aEEG review (13). When we compare the performance of the 12-epoch window used to generate our AUC curves, we find that it still achieves higher sensitivity than standard manual aEEG while maintaining a moderate false positive rate (Figure 5C).

#### Effect of Seizure Duration on Algorithm Performance

Brief seizure durations have been cited as a major cause of poor manual detection by clinicians (10,13). To determine if seizure duration also influences algorithm performance, AUC scores were plotted as a function of average seizure duration for the 39 seizure patients (Figure 6). For all three classifiers, AUC scores tended to increase with seizure duration (Figure 6); the patient with the longest average seizure duration of 918 seconds had AUC scores of 0.90, 0.94, and 0.92 for the RF, SVM and ANN classifiers respectively. In contrast, the patient with the lowest average seizure duration of 10 seconds had AUC scores of 0.86, 0.31, and 0.84. AUC was correlated with seizure duration in the RF (Figure 6A, p=0.028, Spearman Rank Correlation) and ANN (Figure 6C, p=0.0013) but not the SVM classifier (Figure 6B, p=0.14).

**Figure 6.**
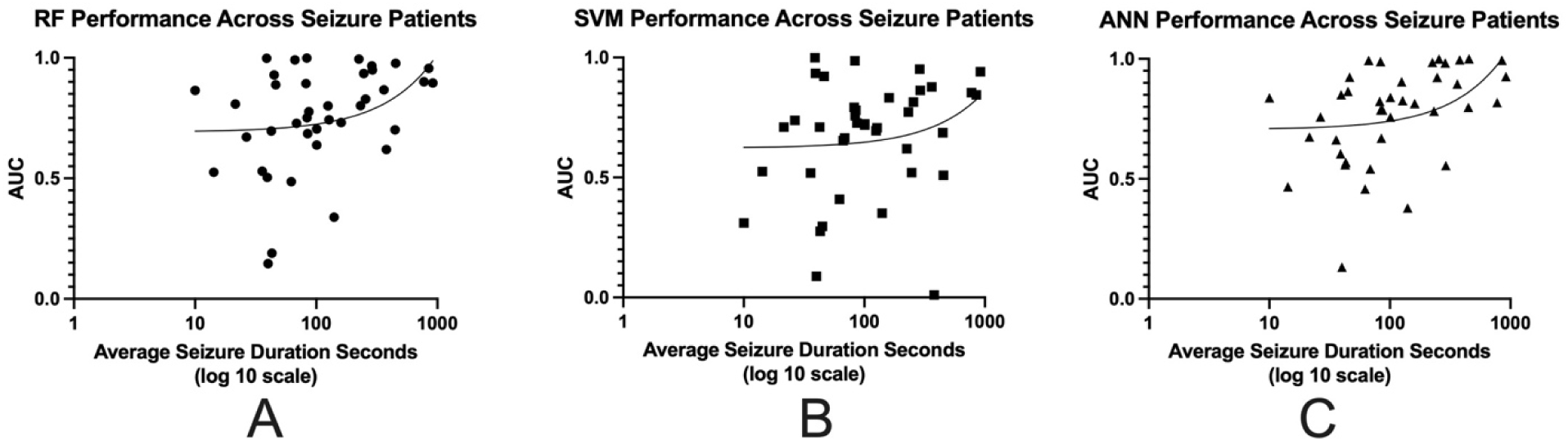
Algorithm performance was evaluated using Spearman rank correlation to assess the relationship between AUC and average seizure duration for the 39 patients with seizures. A. The RF classifier shows increasing AUC values with longer average seizure duration and demonstrates a significant positive correlation (p = 0.028). B. The SVM classifier shows the same upward trend, although the correlation is not significant (p = 0.14). C. The ANN classifier also shows higher AUC values with longer seizure duration and demonstrates a significant correlation (p = 0.0013).

## Discussion

### Clinical Rationale

We conducted a proof-of-concept study to determine whether standard clinical features from two widely used neonatal qEEG systems, aEEG and CSA, could be combined to detect seizures using supervised machine-learning methods. This is the first study to integrate clinical features from both modalities, each of which is already part of routine bedside monitoring. Prior work, such as Lommen et al. (2007), extracted aEEG features but evaluated performance against seizures identified on clinical aEEG rather than EEG, the gold standard (39). Our approach instead uses EEG-verified seizures and relies on features that map directly onto patterns clinicians already recognize, which provides both interpretable detection and insight into which qEEG characteristics best distinguish seizures from background.

Combining aEEG and CSA reflects how clinicians interpret qEEG in practice. aEEG summarizes amplitude-based background and margin trends, while CSA highlights frequency-based changes that often precede or accompany electrographic seizures (9,17,40). Clinicians routinely evaluate both trends alongside the raw EEG for expedited manual review. Incorporating both into a single algorithm therefore mirrors real-world interpretation and provides a clinically grounded, integrated analysis of qEEG features. Supervised machine-learning classifiers were selected because seizures produce nonlinear, multivariate changes across amplitude and spectral domains that are not well captured by simple threshold-based rules used in manual aEEG or CSA review. RF, SVM, and ANN models can learn these nonlinear relationships while still using clinically meaningful features like the aEEG margins or spectral power.

### Limitations

#### Sensitivity vs False Positive Rate Tradeoff in Post-Processing

Post-processing the seizure-probability scores with a moving-average smoothing window created a clear tradeoff between sensitivity and false positive rate. As shown in Figure 5, longer smoothing windows reduced the false positive rate (Figure 5A) but also lowered the percentage of seizures detected (Figure 5B). This occurs because smoothing reduces brief, spurious increases in the algorithm’s seizure-probability estimate during true non-seizure epochs, which helps lower the false positive rate. However, it also reduces short seizure-related changes during true seizure epochs, which lowers sensitivity.

For our AUC assessments, we selected a 12-epoch smoothing window because it corresponds to the average seizure duration in this dataset, which was approximately 115 seconds. At this window length, our algorithm detected a higher fraction of seizures than standard manual aEEG, which typically identifies only about one quarter of electrographic seizures (13,41). These results show that the chosen smoothing window provides a practical balance between limiting false alarms and maintaining adequate seizure detection.

#### Effect of HIE on Algorithm Performance

HIE is the most common cause of neonatal seizures (42), so we examined whether HIE status affected our algorithm’s performance. In this dataset, 56% of seizure patients had HIE (25 of 39), making it the largest subgroup. The classifiers performed well overall (accuracy >0.92), but accuracy was lower in HIE infants than in those without HIE (0.92–0.93 vs. 1.0). This difference likely reflects the imbalance between seizure and non-seizure epochs: there are far more non-seizure epochs overall, making it easier for the classifiers to learn non-seizure patterns than seizure patterns. Since most seizure-positive infants had HIE, the models saw fewer seizure epochs per HIE patient, which lowered per-patient accuracy. Our analysis was limited to comparing HIE and non-HIE groups within this dataset. Future studies using an independent HIE-only cohort will be important for assessing generalizability.

#### Effect of Seizure Duration on Algorithm Performance

Seizure duration is a key factor in detectability (13,43), and seizures under 30 seconds are frequently missed on manual aEEG (10,13). This limitation stems from the heavy time-compression used to generate the bedside aEEG trace: when several minutes of EEG are condensed into a narrow display, brief events are smoothed out and become difficult to distinguish from background activity. Because most neonatal seizures last less than five minutes (42), many short seizures are effectively lost within this compression. Consistent with these clinical observations, all three classifiers in our study detected a higher fraction of longer-duration seizures than shorter ones (Figure 6).

#### Why Our Epoch-Level Metric Likely Underestimates Detection

We used a strict epoch-level definition of true positives and true negatives when evaluating classifier performance (13,41). In this approach, a seizure was counted as missed if any epoch within the seizure event was not labeled as seizure. This approach likely underestimates clinical performance because partial detections would still alert clinicians in actual clinical practice. Prior studies have instead used a Good Detection criterion (16,20), in which a seizure is considered captured if at least one epoch within the event is flagged as seizure. Using Good Detection would increase sensitivity without changing the false positive rate and better reflects how clinicians interpret qEEG trends. Incorporating this criterion in future validation studies will provide a more accurate representation of clinical utility.

#### Benefits and Limitations of Centroparietal qEEG Features

Both aEEG and CSA remain widely used bedside tools because they are familiar to clinicians and easy to implement using a limited electrode montage. However, this limited electrode setup reduces spatial resolution and can obscure focal seizures, which is why qEEG trends are often used as adjuncts rather than replacements for continuous EEG review (14).

These practical and spatial constraints also shaped our computational approach. Since bedside aEEG and CSA rely on a limited montage, we intentionally designed our feature set to mirror the limited array. Maintaining this alignment ensures that the algorithm remains simple, interpretable, and grounded in the same physiologic patterns clinicians already use when reviewing qEEG trends.

We selected channels C3-P3 and C4-P4 because they are the recommended electrode pairs used for bedside aEEG since most neonatal seizures arise in the central region (6,13,44,45). These centroparietal channels also provide reliable coverage of the perirolandic cortex, an area where many neonatal seizures originate (46). They are also less susceptible to artifact than frontal or temporal electrodes (44). Despite these advantages, relying solely on these channels increases the likelihood of missing seizures that arise from frontal, temporal, or occipital regions, even if they only account for a minority of neonatal seizures (2).

#### Need for Independent Cohort Evaluation

This study was designed as a proof-of-concept evaluation of whether clinically relevant aEEG and CSA features can support automated seizure detection. All model development was performed using patient-level three-fold cross-validation, which ensured that no patient contributed data to both the training and testing sets within the same fold. This approach reduces information leakage and prevents the models from learning patient-specific patterns. However, because the entire analysis was completed within a single publicly available dataset, the results reflect performance in this specific cohort rather than generalizable accuracy across NICU populations.

Neonatal EEG varies across centers, equipment, and clinical conditions, which can affect how well a model trained on one dataset performs on another. To determine whether this approach generalizes beyond the Helsinki cohort, future work should evaluate the feature set and modeling framework on an independent dataset that captures a broader range of neonatal EEG backgrounds. Such studies will clarify algorithm robustness and help establish whether a combined aEEG-CSA approach can support broader clinical use.

## Conclusion

This proof-of-concept study shows that combining amplitude-integrated EEG and compressed spectral array features with supervised machine-learning models can help identify electrographic seizure patterns in neonates using clinically interpretable qEEG features. The Random Forest, Support Vector Machine, and Artificial Neural Network classifiers each captured physiologic changes associated with seizure evolution, with the Random Forest model performing most consistently across patients. The most informative features reflected both amplitude-based and frequency-based dynamics, underscoring the complementary value of integrating aEEG and CSA trends.

Our algorithm’s performance varied with seizure duration and etiology, with infants who had hypoxic-ischemic encephalopathy and those with shorter seizures showing lower detection accuracy. Despite these constraints, the model still identified a substantial proportion of electrographic seizures using only standard bedside qEEG features. These results demonstrate the feasibility of an interpretable aEEG-CSA framework for neonatal seizure detection and highlight the need for future studies incorporating broader electrode coverage, larger cohorts, and external validation.

## Data Availability

All data produced in the present study are available upon reasonable request to the authors

## Clinical Trial Number

Not applicable.

## Declarations

### Ethics approval and consent to participate

This study was conducted ethically in compliance with the World Medical Association Declaration of Helsinki. Written consent was not required for this retrospective study.

### Consent for publication

All authors have provided consent for the publication of this manuscript.

### Availability of data and material

All data generated and evaluated for this study are included in this article. Further enquiries may be directed to the corresponding author.

### Competing interests

The authors have no relevant conflicts of interest.

### Funding

This study was funded by the National Institute of General Medical Sciences T32 training grant.

### Authors’ contributions

SE, JH, HD, and NPI conceptualized the article. SE conducted aEEG feature extraction. SE, NPI, and JH evaluated aEEG features. SE conducted CSA feature extraction. SE, NPI, and HD evaluated CSA features. SE implemented the machine learning algorithms, and BBJ ran and evaluated the machine learning algorithms. SE, NPI, and BBJ performed statistical analysis. SE and NPI drafted the article. SE, NPI, BBJ, and JH edited the article. SE, NPI, BBJ, JH, and HD proofread the article.

## Acknowledgments

We are grateful to the University of Chicago’s department of pediatrics section of neurology, the pediatric EEG technical staff at Comer Children’s Hospital, and the University of Chicago’s department of medicine.

## Supplemental Table

**Supplemental Table 1:** Documented parameter settings used to generate the two aEEG versions (aEEG A and aEEG B).

| *aEEG*<br>*Version* | *Reference* | *Asymmetric Bandpass Filter*<br>*(2-15Hz)* | *Enveloping Filter* |
| --- | --- | --- | --- |
| *aEEG A* | Werther et al<br>2018 | Linear Phase FIR filter<br>(Parks-McClellan) Filter 2 | Rectangular Moving<br>Average Filter (3-second<br>window) |
| *aEEG B* | Werther et al<br>2018 | Linear Phase FIR filter<br>(Parks-McClellan) Filter 5 | Rectangular Moving<br>Average Filter (0.5-second window) |

## Notes

### Competing Interest Statement

The authors have declared no competing interest.

### Author Declarations

This study used data from publicly available neonatal EEG dataset published by Stevenson et al 2019 in Nature Scientific Data.

